# Gaze, Visual, Myoelectric, and Inertial Data of Grasps for Intelligent Prosthetics

**DOI:** 10.1101/19010199

**Authors:** Matteo Cognolato, Arjan Gijsberts, Valentina Gregori, Gianluca Saetta, Katia Giacomino, Anne-Gabrielle Mittaz Hager, Andrea Gigli, Diego Faccio, Cesare Tiengo, Franco Bassetto, Barbara Caputo, Peter Brugger, Manfredo Atzori, Henning Müller

## Abstract

Hand amputation is a highly disabling event, having severe physical and psychological repercussions on a person’s life. Despite extensive efforts devoted to restoring the missing functionality via dexterous myoelectric hand prostheses, natural and robust control usable in everyday life is still challenging. Novel techniques have been proposed to overcome the current limitations, among which the fusion of surface electromyography with other sources of contextual information. We present a dataset to investigate the inclusion of eye tracking and first person video to provide more stable intent recognition for prosthetic control. This multimodal dataset contains surface electromyography and accelerometry of the forearm, and gaze, first person video, and inertial measurements of the head recorded from 15 transradial amputees and 30 able-bodied subjects performing grasping tasks. Besides the intended application for upper-limb prosthetics, we also foresee uses for this dataset to study eye-hand coordination in the context of psychophysics, neuroscience, and assistive robotics.

## Background & Summary

The human hand is said to separate man from primates. We use it to sense and interact with the environment, and it has an essential role in social interactions and communication. Losing a hand therefore has severe physical and psychological consequences on a person’s life. Myoelectric hand prostheses have enabled amputees to restore some level of the missing functionality. In their conventional form, these use surface electromyography (sEMG) from an antagonist pair of muscles to open and close a simple gripper. A large number of pattern recognition (PR) approaches have been developed over the years aiming to increase the functionality and control of more dexterous prostheses [51, 10, 27]. Despite these efforts, PR-based prostheses have not managed to deliver their full potential in practical clinical applications [42, 47]. One of the primary causes is that they are sensitive to the variability of sEMG, limiting the robustness and reliability in long-term practical applications [45, 38, 43, 10, 22, 50, 21, 14]. This limitation is inherent in the use of sEMG as a control modality, since myoelectric signals are known to depend on factors such as electrode displacement, inconsistency of the skin-electrode interface, changes in arm position, and characteristics of the amputation.

Several techniques have been proposed to address this variability and thus improve the stability of the prosthetic control [10, 22, 2, 24]. One approach relies on fusing sEMG with complementary sources of information, such as gaze and computer vision [9, 13, 17]. Studies on eye-hand coordination have in fact shown that gaze typically anticipates and guides grasping and manipulation [30, 36, 18]. In other words, humans will look at an object they intend to grasp before executing the movement itself. Gaze behavior therefore holds valuable information for recognizing not only the intention to grasp an object, but also to identify *which* object the person intends to grasp. The set of likely grasps can then be estimated based on the size, shape, and affordances of this object. For large objects, this may be further refined by considering the exact part or side of the object on which the person fixates.

There are two compelling motivations for using gaze behavior in the context of upper limb prosthetics. The first is that gaze behavior is not necessarily affected by the amputation. Any information on muscular activation recorded from the residual limb, such as sEMG, is inherently degraded due to muscular reorganization after the amputation and subsequent atrophy due to reduced use. The second advantage is that integrating some level of autonomy in a prosthesis can lower the physical and psychological burden placed on its user. Fatigue is one of the causes of variability in sEMG data, so reducing it can help to stabilize control.

This paper presents the MeganePro dataset 1 (MDS1) acquired during the MeganePro project, which was designed based on the experience gained during the NinaPro project [4]. The goals of the MeganePro project were to investigate the use of gaze and computer vision to improve the control of myoelectric hand prostheses for transradial amputees, and to better understand the neurocognitive effects of amputation. This dataset was acquired to explore the first goal: it contains multimodal data from 15 transradial amputees and 30 able-bodied subjects while involved in grasping and manipulating objects using 10 common grasps. Throughout the exercise, we acquired sEMG and accelerometry using twelve electrodes on the forearm. Gaze, first person video, and inertial measurements of the head were recorded using eye tracking glasses.

In the following sections, we will describe the experimental protocol, the processing procedures, and the resulting data records. Although prosthetic control was our motivation for acquiring this dataset, it is our belief that the data will find broader applications. For instance, scientists working in neuroscience, psychophysics, or rehabilitation robotics may use it to study gaze and manipulation independently as well as via their coordination. In particular, the data allow direct comparison of able-bodied subjects and transradial amputees in an identical experimental setting. As such, it may contribute not only to develop better prostheses, but also a better understanding of human behavior and the implications of amputation.

## Methods

The acquisition of the dataset consisted of defining an experimental protocol and determining the requirements in terms of devices and software. A general overview of the setup and protocol is shown in Figure 1. Once the ethical approval was obtained, we proceeded with subject recruitment. In the following, all these phases will be described in detail.

**Figure 1:**
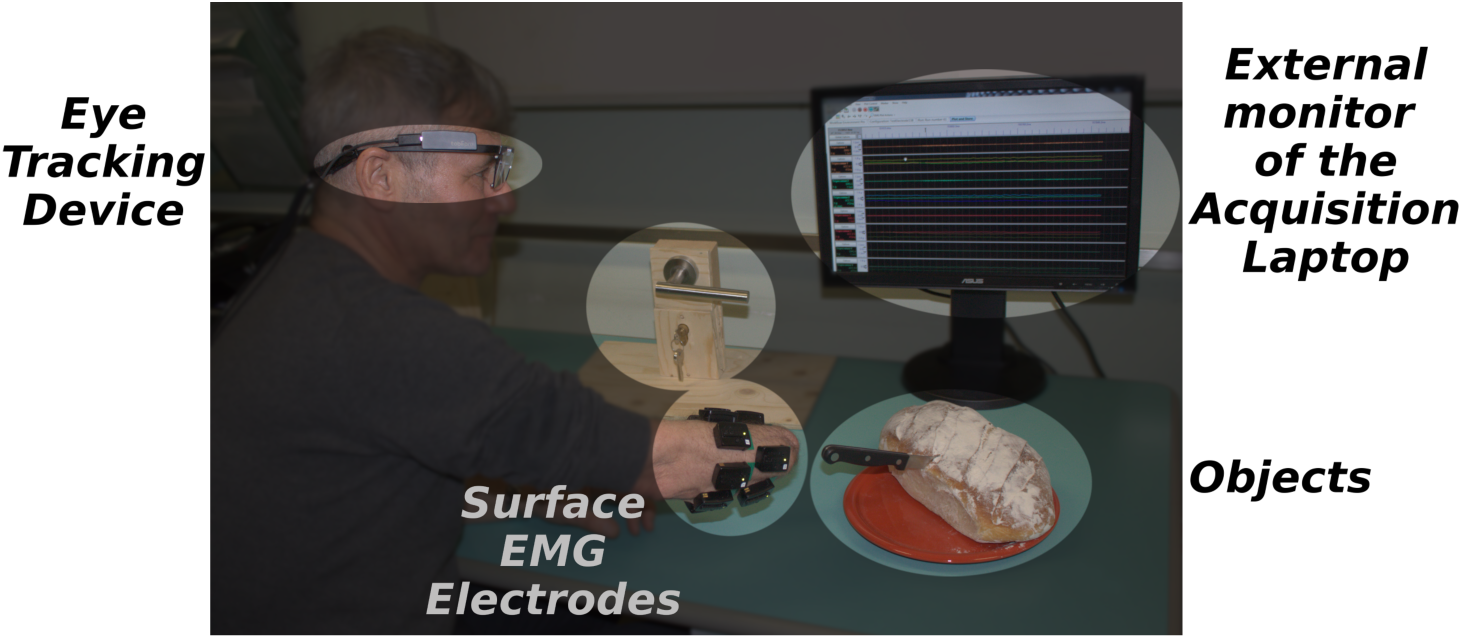
Overall view of the acquisition setup.

### Ethical Requirements

The experiment was designed and conducted in accordance with the principles expressed in the Declaration of Helsinki. Ethical approval for our study was requested to and approved by the Ethics Commission of the canton of Valais in Switzerland and by the Ethics Commission of the Province of Padova in Italy. Prior to the experiment, each subject was given a detailed written and oral explanation of the experimental setup and protocol. They were then required to give informed consent to participate in the research study.

### Subject Recruitment

A total of 15 transradial amputees and 30 able-bodied subjects were recruited for this study. The former group consists of 13 male and 2 female (13.3%) participants with an average age of (47.13 ± 14.16) years. As seen in Table 1, among them there are several causes for amputation and different preferences with respect of prosthetic use. To remove possible confounding variables, we recruited za control group of able-bodied participants that matched the former group as much as possible in terms of age and gender. This group is composed of 27 male and 3 female (10.0%) subjects, with an average age of (46.63 ± 15.11) years.

**Table 1:**
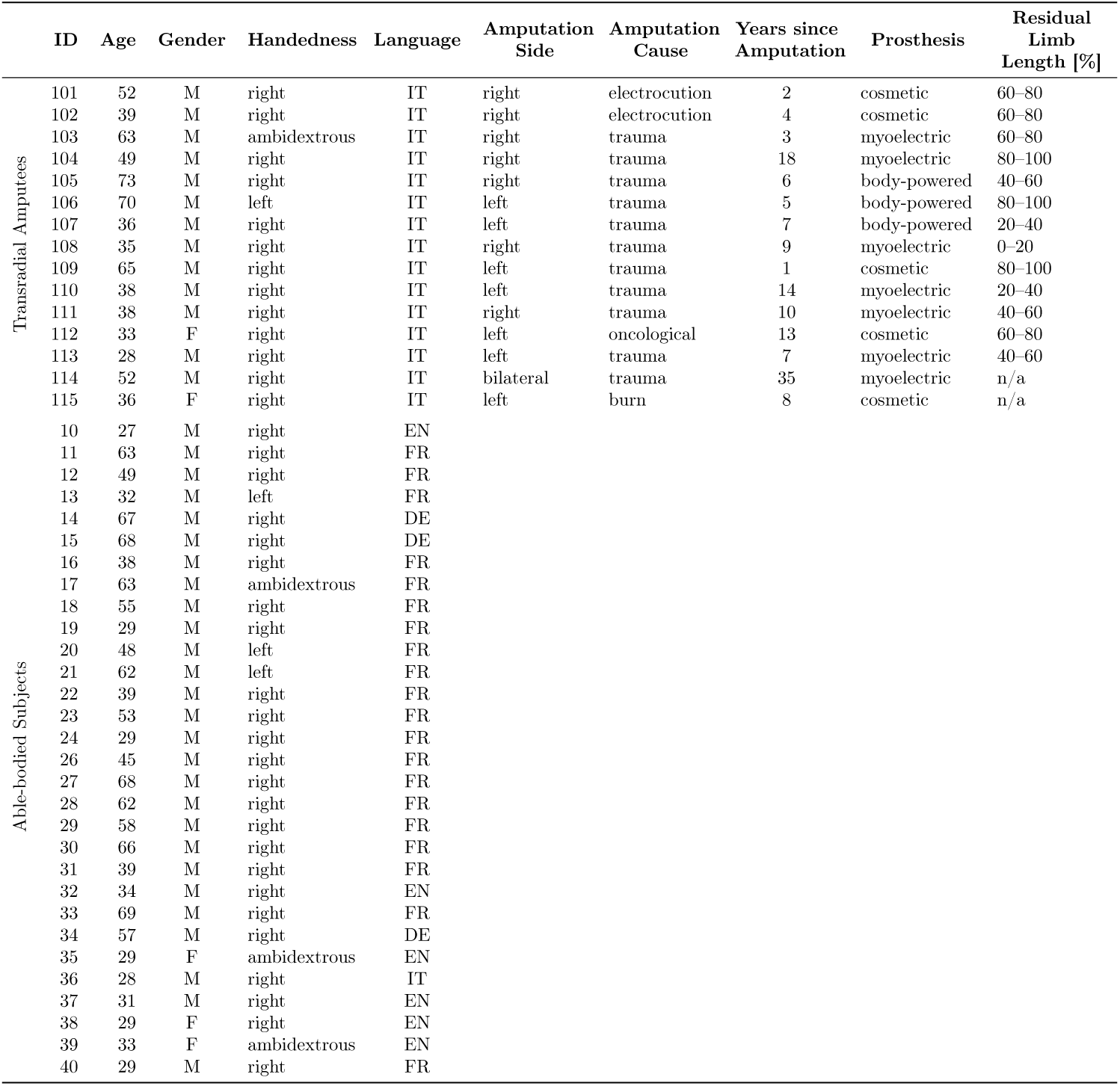
Participant characteristics. The table reports the ID of the subject in the dataset, their age, their gender and their handedness for all the subjects. Clinical parameters about the amputation(s) are also reported for the transradial amputees. The rightmost column indicates the relative length of the residual limb with respect to the contralateral limb.

### Acquisition Setup

We designed an acquisition setup that allowed us to perform eye tracking and to acquire sEMG from the forearm, while leaving the subjects as free as possible in their movements. Custom developed software interfaced with the acquisition devices and provided stimuli instructing the subjects when to grasp which object.

#### Sensors

The electrical activity of the forearm muscles was recorded with a Delsys Trigno Wireless sEMG System (Delsys Inc., USA)^1^. This system consists of sEMG electrodes with an inter-sensor latency lower than 500 µs and a signal baseline noise lower than 750 nV Root Mean Square (RMS). In addition, a three-axial accelerometer is embedded in each electrode. Up to 16 electrodes communicate wirelessly with the base station, which is connected through a USB 2.0 cable to the acquisition laptop. The sEMG data are sampled at 1926 Hz and the accelerometer at 148 Hz.

The gaze behavior and first person video were recorded with the Tobii Pro Glasses 2 (Tobii AB, Sweden)^2^. The head unit of this device is similar to regular eyeglasses and equipped with four eye cameras recording the eye movement, a camera capturing the scene in front of the subject, an Inertial Measurement Unit (IMU), and a microphone. It weighs only 45 g and corrective lenses can be applied for subjects with visual deficit. The eyes are tracked using corneal reflection and dark pupil methods with automatic parallax and slippage compensation. This eye tracking data are sampled at 100 Hz with a theoretical accuracy and precision of 0.5° and 0.3° RMS. Video with Full HD resolution (1920 px × 1080 px) is recorded at 25 frames per second by the scene camera with a horizontal and vertical field of view of approximately 82° and 52°. The head unit is connected via a cable to a portable recording unit that is responsible for transmitting the data wirelessly and storing them on an memory card. A rechargeable and replaceable Li-ion battery powers both head and recording units and provides a maximum recording time of approximately 120 min. The system can quickly and easily be calibrated with a single point calibration procedure.

#### Acquisition Software

The role of the acquisition software is to simultaneously acquire and store the data from all sensor devices, and to guide the subject through the exercises. To ensure high performance and low latency, the application was developed in C++ and based on the multithreaded producer-consumer pattern as implemented in CEINMS [40]. More specifically, for each data source there is dedicated producer thread responsible for acquiring its data, assign a high-resolution timestamp, and then store these in a queue. Per queue there is at least one consumer thread that stores available data to a file. The advantage of this architecture is to uncouple the data acquisition from I/O latencies when saving the data to persistent storage.

A Graphical User Interface (GUI) developed in Qt5^3^ was added to the application to demonstrate using videos and images how to perform the exercises. In previous studies, such as NinaPro [3], subjects simply had to mimic a grasp movement that was shown in a video. This approach is not compatible with the present study, since it would have influenced the subject’s gaze behavior. We therefore opted to avoid the need for visual attention and to instruct the subjects via vocal instructions during the real exercise. These instructions were automatically synthesized by a text-to-speech engine, which allowed us to prepare instructions in English, Italian, French, and German. At the start of the exercise subjects could choose the language they were most comfortable with.

#### Grasp Types and Objects

A total of ten grasp types were selected for our exercise from well-known hand taxonomies [16, 46, 15, 23] based on their relevance for Activities of Daily Living (ADLs) [8]. These grasp types were then matched with three household objects each that would regularly be manipulated using a given grasp. An overview of the ten grasp types and eighteen objects is given in Table 2. When possible, these pairings were chosen to obtain a many-to-many relationship between objects and grasp types; that is, a grasp could be used with multiple objects and vice versa. This condition helped to limit the possibility that the mere presence of an object would be sufficient to unequivocally determine the grasp type.

**Table 2:** Overview of the grasp types and objects for the *static* condition of the exercise. The columns indicate the ID and name of the grasp as commonly reported literature [23, 16], the ID and name of the object, and in some cases a further refinement indicating the ID and name of the part of the object that was involved in the grasping. The fourth column reports the vocal command given to the subject.

| Grasp |  | Object |  | Object Part | Vocal Instruction |
| --- | --- | --- | --- | --- | --- |
| 1 | medium wrap | 1 | bottle | 1 bottle | take the bottle |
|  |  | 2 | can | 2 can | can |
|  |  | 3 | door handle | 3 door handle | door handle |
| 2 | lateral | 4 | mug | 23 handle | mug |
|  |  | 5 | key | 5 key | key |
|  |  | 24 | pencil case | 6 zip | zip |
| 3 | parallel extension | 7 | plate | 7 plate | plate |
|  |  | 8 | book | 8 book | book |
|  |  | 9 | drawer | 9 drawer | drawer |
| 4 | tripod grasp | 1 | bottle | 10 cap | cap of the bottle |
|  |  | 4 | mug | 4 mug | mug |
|  |  | 9 | drawer | 11 knob | knob of the drawer |
| 5 | power sphere | 12 | ball | 12 ball | ball |
|  |  | 13 | bulb | 13 bulb | light bulb |
|  |  | 5 | key | 5 key | keys |
| 6 | precision disk | 15 | jar | 26 lid | jar |
|  |  | 13 | bulb | 13 bulb | light bulb |
|  |  | 12 | ball | 12 ball | ball |
| 7 | prismatic pinch | 16 | clothespin | 16 clothespin | clothespin |
|  |  | 5 | key | 27 keyring | keys |
|  |  | 2 | can | 25 pull tab | can |
| 8 | index finger extension | 21 | remote | 17 button | point at a button of the remote |
|  |  | 18 | knife | 18 knife | take the knife |
|  |  | 19 | fork | 19 fork | fork |
| 9 | adducted thumb | 20 | screwdriver | 20 screwdriver | screwdriver |
|  |  | 21 | remote | 21 remote | remote |
|  |  | 22 | wrench | 22 wrench | wrench |
| 10 | prismatic four finger | 18 | knife | 18 knife | knife |
|  |  | 19 | fork | 19 fork | fork |
|  |  | 22 | wrench | 22 wrench | wrench |

In the case that an object is not usually found on a table (e.g., a door handle or a door lock), a custom made support was created. To avoid complications during the exercise, the key, bulb, lid of the jar, and the screw used with the screwdriver were modified such that they could not be completely removed from the support. Furthermore, the pull tab of the can was bent to facilitate the execution of the grasp.

During the exercise, at least five objects were simultaneously placed in the scene with a predefined position. The presence of multiple objects in front of the subjects helped to simulate a real environment, while a standardized object arrangement minimized the possibility of an error during the placement of the objects. In addition, these objects were spread throughout the subject’s reachable workspace so that grasps had to be performed while reaching in various directions.

### Acquisition Protocol

After the ethical requirements were fulfilled, the subject’s personal data, such as age, gender, height, weight, and handedness were collected. The amputees were asked additional information regarding the amputation, such as cause, type, years since amputation, and prosthesis use. A summary of this information is reported in Table 1.

The subjects were asked to sit with the forearm comfortably leaning on a desk. The skin was cleaned with isopropyl alcohol and twelve sEMG electrodes were placed in two arrays around the right forearm or residual limb. An array of eight electrodes was placed equidistantly around the forearm, starting at the radio-humeral joint and moving in the direction of pronation. A second array was located approximately 45 mm more distally and aligned with the gaps between electrodes one and two, three and four, and so on (see Figure 1). In addition to the Trigno-specific adhesive strips, a latex-free elastic band was wrapped around the electrodes to assure a good and reliable interface with the skin.

The subject was then asked to wear the Tobii Pro glasses, where we made sure to choose the nose pad that aligned the eye tracking cameras’ appropriately. Once the subject felt ready, the Tobii Pro glasses were calibrated using the built-in one point target calibration procedure and, in case of success, the subject was asked to perform a calibration assessment. This assessment consisted in asking the subject to fixate a black cross against a green background that was displayed on a monitor at a distance of about 1.3 m. This cross remained fixed for 3 s in the same location before alternating to another one out of five positions in total.

The actual exercise consisted of repeatedly executing the grasps on the set of corresponding objects, as described in Table 2. In the first part, which we will refer to as *static*, subjects were requested to grasp the objects without actually moving or lifting them. Prior to executing a grasp, videos in first and third person perspectives demonstrated the grasp and the three corresponding objects. The subjects were however instructed to execute the movements as naturally as possible, rather than attempting to mimic the exact kinematics of the demonstration videos. During these videos, the subject was free to decide whether to perform or not the grasps to get confident about the exercise. Once the grasp and objects had been shown, the subject had to repeat each combination four times while seated and then another four times while standing. This change in position was intended to include variability in the limb position in the data. The duration of the movement and rest periods depends on the selected language for the vocal instructions. A grasp interval lasted approximately 5.2 s, 5.7 s, 5.9 s, and 6.0 s for English, Italian, French, and German. A rest period followed for about 4.1 s, 4.7 s, 4.7 s, and 4.7 s for English, Italian, French, and German. The exact order of the objects within each repetition was randomized to avoid learning and habituation effects. A vocal command guided the subject through the exercise, indicating which object to grasp, when to return to the rest position, and when to stand up or sit. These instructions were accompanied by a static image of the current grasp type, which was intended as an undistracting reminder of the stimulus. When finishing a grasp series, an experimenter changed the object scene when necessary before resuming with the next grasp.

The *static* part of the exercise was followed by a *dynamic* one. This second part followed the same structure as the first, but in this case the grasp had to be used to perform a functional task with an object. The motivation for this variation was again to introduce variability in the data, but this time by means of a dynamic, goal-oriented movement. As can be seen in Table 3, these functional movements reflect common activities with the combination of grasp type and object, such as drinking from a can held with a medium wrap. To limit the total duration of the exercise, the ten grasps were only combined with two objects each and executed either standing or seated. This position was chosen based on how the action would usually be performed in real life, for instance a door is more frequently opened while standing.

**Table 3:** Overview of grasp types and objects for the *dynamic* condition of the exercise. The first three columns provide information as described in Table 2. The rightmost column indicates whether the movement was executed while seated or standing.

| Grasp | Object | Object Part | Vocal Instruction | Position |
| --- | --- | --- | --- | --- |
| 1 medium wrap | 2 can | 2 can | drink from the can | standing |
|  | 3 door handle | 3 door handle | open and close the door handle |  |
| 2 lateral | 5 key | 5 key | turn the key in the lock | standing |
|  | 24 pencil case | 6 zip | open and close the pencil case |  |
| 3 parallel extension | 7 plate | 7 plate | lift the plate | standing |
|  | 8 book | 8 book | lift the book |  |
| 4 tripod grasp | 1 bottle | 10 cap | open and close the cap of the bottle | standing |
|  | 9 drawer | 11 knob | open and close the drawer |  |
| 5 power sphere | 12 ball | 12 ball | move the ball to the right and back | standing |
|  | 5 key | 5 key | move the keys forwards and backwards |  |
| 6 precision disk | 15 jar | 26 lid | open and close the lid of jar | seated |
|  | 13 bulb | 13 bulb | screw and unscrew the light bulb |  |
| 7 prismatic pinch | 16 clothespin | 16 clothespin | squeeze the clothespin | seated |
|  | 5 key | 27 keyring | move the keys forwards and backwards |  |
| 8 index finger extension | 21 remote | 17 button | press a button on the remote control | seated |
|  | 18 knife | 18 knife | cut bread with the knife |  |
| 9 adducted thumb | 20 screwdriver | 20 screwdriver | turn the screwdriver | seated |
|  | 22 wrench | 22 wrench | move the wrench to the right and back |  |
| 10 prismatic four finger | 18 knife | 18 knife | move the knife forwards and backwards | seated |
|  | 19 fork | 19 fork | move the fork to the right and back |  |

After the exercise finished the amputated subjects underwent three more exercises and a comprehensive questionnaire to investigate the neurocognitive effects of the amputation. The data and results of those exercises and the questionnaire will be published separately.

### Post-Processing

A number of processing steps were applied to the raw data acquired with the protocol described above. The objective of these steps was to sanitize the data, synchronize all modalities, and remove identifying information from the videos. In the following we describe all procedures in detail.

#### Timestamp Correction

Due to an unfortunate implementation error, during a number of acquisitions the modalities were assigned timestamps from individual clocks. To unify all timestamps in a shared clock, the offset of all clocks was estimated and corrected with respect to the clock of the sEMG modality using statistics of their relative timing collected during trial acquisitions. Validations on the remaining unaffected acquisitions confirm that the maximum deviation of our estimate from the ground truth is less than 12 ms.

#### sEMG and Accelerometer Data

For computational efficiency, the sEMG and accelerometer streams from the Delsys Trigno device were acquired and timestamped in batches. During post-processing, individual timestamps were assigned to each sample via piecewise linear interpolation. A new piece is created if the linear model would result in a deviation of more than 100 ms, which may happen if the fit is skewed due to missing or delayed data.

For the sEMG data, we furthermore filtered outliers by replacing samples that exceeded 30 standard deviations from the mean within a sliding window of 1 s with the preceding sample. The signals were subsequently filtered for power-line interference at 50 Hz (and its harmonics) using a Hampel filter, which interpolates the spectrum in processing windows only when it detects a clear peak [1]. Contrary to the more common notch filter, this method does not affect the spectrum if there is no interference.

#### Gaze Data

The data from the Tobii Pro glasses were acquired as individually timestamped JavaScript Object Notation (JSON) messages. During post-processing, these messages were decoded and separated based on their type. Messages that arrived out-of-order were filtered and the resulting set of messages was used to determine the skew and offset between the computer clock and the one of the Tobii Pro glasses. This routine removes the constant part of the transmission delay as much as possible, while avoiding the possibility to antedate any events. The messages that relate directly to gaze information, such as gaze points, pupil diameter and so on, were then grouped together based on their timestamps.

#### Stimulus

The text-to-speech engine that was used to give vocal instructions introduced noticeable delays in the corresponding changes of the stimulus. We measured these delays for all sentences and languages, and moved the stimulus changes forward by these amounts during post-processing. Furthermore, small refinements were made to the *object* column, whereas its more specific *object-part* counterpart was calculated based on a fixed mapping from the original stimulus information.

#### Synchronization

For the standard data records (see following section) all modalities were resampled at the original 1926 Hz sampling rate of the sEMG stream. For real-valued signals, this was done using linear interpolation, while for discrete signals we used nearest-neighbor interpolation. The signals that indicate the time and index of the MP4 video were handled separately using a custom routine, since they require to identify the exact change-point where one video transitions to the next.

#### Concatenation

The *static* and *dynamic* parts of the protocol were acquired independently and therefore produced separate sets of raw acquisition files. Furthermore, our acquisition protocol and software allowed to interrupt and resume the acquisition, either at request of the subject or to handle technical problems. After applying the previous processing steps to the individual acquisition segments, they were concatenated to obtain the standard data record. During this merging, we incremented the timestamps and video counter to ensure that they are monotonically increasing. Furthermore, if part of the protocol was repeated when resuming the acquisition we took care to insert the novel segment at exactly the right place to avoid duplicate data.

#### Relabeling

The response of the subject and therefore the sEMG activation may not be aligned perfectly with the stimulus. As a consequence, the stimulus labels around the on- or offset of a grasp movement may be incorrect, resulting in an undue reduction in recognition performance. We addressed this shortcoming by realigning the stimulus boundaries with the procedure described by Kuzborskij et al. [35]. In short, this method optimizes the log-likelihood of a rest-grasp-rest sequence on the whitened sEMG data within a feasible window that spans from 1 s before until 2 s after the original grasp stimulus. As opposed to the uniform prior used in the earlier method, we instead adopted a smoothed variant of the original stimulus label as prior with *p* = 0.6. The recalculated stimulus boundaries have been saved in addition to the original ones.

#### Removing Identifying Information

All videos were checked manually for identifying information of anyone other than the experimenters. The segments of video that were marked as privacy-sensitive were subsequently anonymized with a Gaussian blur. In this procedure, we took care to re-encode only the private segments and to preserve the exact number and timestamps of all frames. In addition, the audio stream was removed from all videos for privacy reasons.

### Code Availability

The post-processing procedure described above was implemented in Matlab and executed in compiled form with Matlab 2016b. The relabeling procedure instead was implemented in Python and interpreted with Python 3.6. Censoring of the videos was done using a custom Bash script that interfaced with the Ffmpeg 4.1.3 video manipulation tool. The GNU Parallel tool was used to run jobs in parallel [48]. A copy of the code that was used to create the data records from the raw recordings is publicly available [25].

## Data Records

The data that were acquired and processed with the described methodology were stored in Harvard’s Dataverse [12]. For each subject, this repository contains two data files in Matlab format and a series of corresponding videos encoded with MPEG-4 AVC in an MP4 container. The primary data file contains the concatenated sEMG data at their original sampling rate, to which all other modalities were resampled. An exhaustive listing of all the fields in these files is provided in Table 4.

**Table 4:** Fields contained in the standard data record.

| Field | Columns | Units | Description |
| --- | --- | --- | --- |
| grasp | 1 |  | ID of the desired grasp |
| grasprepetition | 1 |  | repetition counter for the desired grasp |
| object | 1 |  | ID of the target object |
| objectpart | 1 |  | ID of the target object part |
| objectrepetition | 1 |  | repetition counter for the target object |
| position | 1 |  | seated or standing position |
| dynamic | 1 |  | static or dynamic grasp indicator |
| regrasp | 1 |  | realigned ID of the desired grasp |
| regrasprepetition | 1 |  | realigned repetition counter for the desired grasp |
| reobject | 1 |  | realigned ID of the target object |
| reobjectpart | 1 |  | realigned ID of the target object part |
| reobjectrepetition | 1 |  | realigned repetition counter for the target object |
| reposition | 1 |  | realigned seated or standing indicator |
| redynamic | 1 |  | realigned dynamic grasp indicator |
| acc | 36 | g | 3-axis acceleration of the 12 electrodes |
| emg | 12 | V | myoelectric activity of the 12 electrodes |
| gazept | 2 |  | 2D gaze point relative to the scene image |
| gazept_invalid | 1 |  | invalidity indicator for "gazept" |
| gazept3D | 3 | mm | 3D gaze point in world coordinates |
| gazept3D_invalid | 1 |  | invalidity indicator for "gazept3D" |
| gazedirectionleft | 3 |  | 3D gaze direction of the left eye |
| gazedirectionleft_invalid | 1 |  | invalidity indicator for "gazedirectionleft" |
| gazedirectionright | 3 |  | 3D gaze direction of the right eye |
| gazedirectionright_invalid | 1 |  | invalidity indicator for "gazedirectionright" |
| pupilcenterleft | 3 | mm | 3D position for the pupil center of the left eye |
| pupilcenterleft_invalid | 1 |  | invalidity indicator for "pupilcenterleft" |
| pupilcenterright | 3 | mm | 3D position for the pupil center of the right eye |
| pupilcenterright_invalid | 1 |  | invalidity indicator for "pupilcenterright" |
| pupildiameterleft | 1 | mm | pupil diameter of the left eye |
| pupildiameterleft_invalid | 1 |  | invalidity indicator for "pupildiameterleft" |
| pupildiameterright | 1 | mm | pupil diameter of the right eye |
| pupildiameterright_invalid | 1 |  | invalidity indicator for "pupildiameterright" |
| tobiacc | 3 | $\text{m s}^{-2}$ | 3-axis acceleration of the Tobii |
| tobiacc_invalid | 1 |  | invalidity indicator for "tobiacc" |
| tobiigr | 3 | $^{\circ} \text{s}^{-1}$ | 3-axis angular velocity of the Tobii |
| tobiigr_invalid | 1 |  | invalidity indicator for "tobiigr" |
| tobiits | 1 | s | timestamp in the Tobii clock |
| vts | 1 | s | MP4 video timestamp |
| mp4videoidx | 1 |  | counter for the MP4 video |
| pts | 1 | s | TS presentation timestamp |
| tspipelineidx | 1 |  | TS pipeline ID |
| tsvideoidx | 1 |  | counter for the TS video |
| ts | 1 | s | timestamp in the computer clock |

Since resampling may not always be desirable due to its impact on the the signal spectrum, we also provide an auxiliary data file with all non-sEMG modalities at their original sampling rate. Each of these includes a *timestamp* column, which can be used to synchronize them with each other or with the sEMG data in the primary data file. The fields and their detailed specification is shown in Table 5.

**Table 5:**
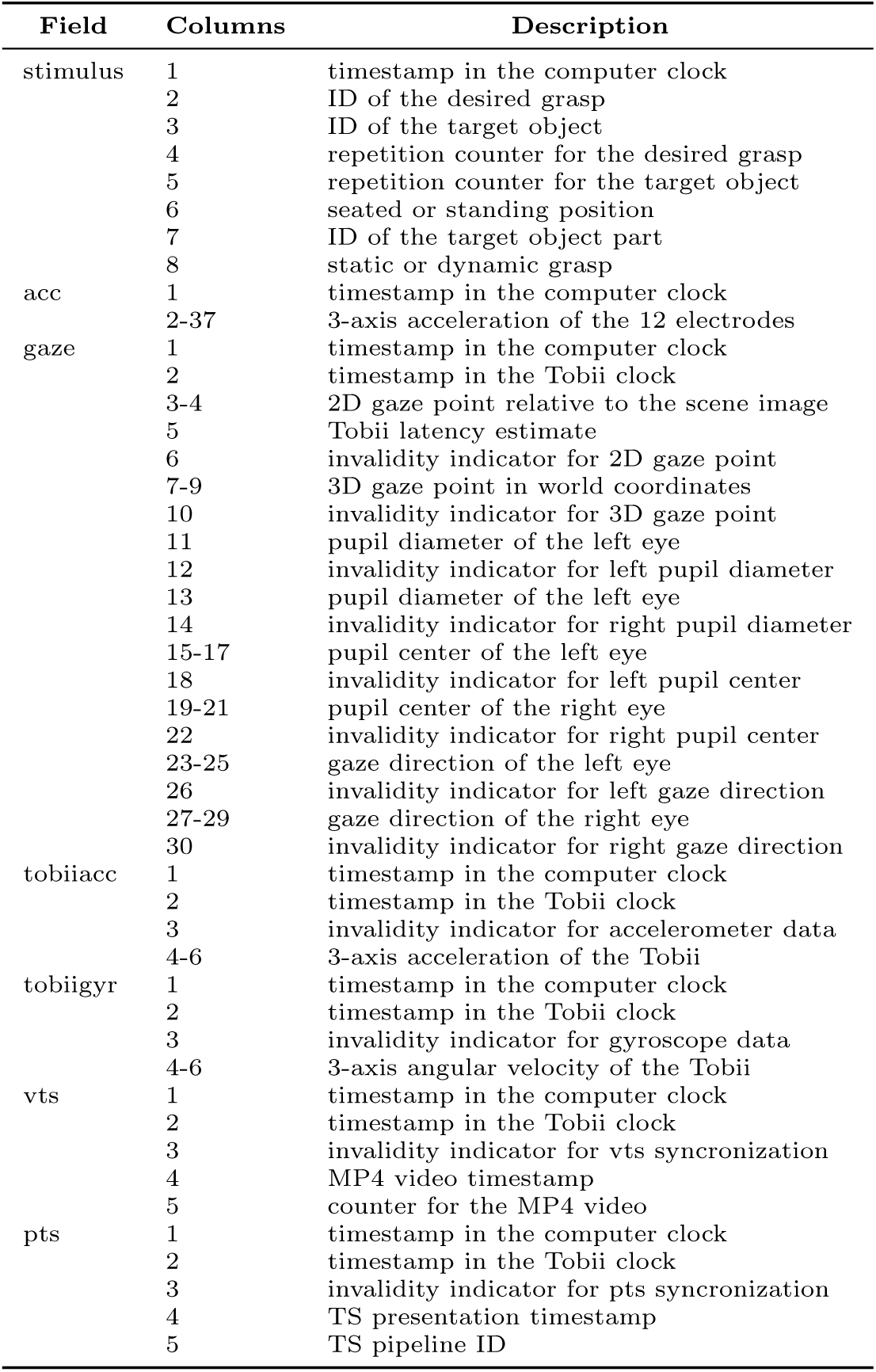
Fields contained in each acquisition segment of the auxiliary data record with original sampling.

## Technical Validation

Our intended purpose for the dataset is to investigate the fusion of sEMG with gaze behavior. In this section we therefore concentrate on validating these two modalities. Some of these analyses are low-level to ensure the quality of the recorded signals, while others are meant to verify that the data can in fact be used for the motivations for which it was created.

### Gaze Data

#### Error Validation

The quality of gaze data primarily depends on the correctness of the initial calibration phase of the Tobii Pro glasses. Validating the effectiveness of this calibration consists in acquiring gaze data while the user is focused on a known target and subsequently comparing the measured gaze location with this ground truth [29, 7]. We used the data recorded during the calibration assessment described in section Acquisition Protocol to evaluate the effectiveness of the calibration as well as possible quality degradation. These data were typically collected at the beginning and end of an exercise. If the exercise was interrupted, the procedure was shown again before resuming it. We determined the ground truth by manually locating the cross position in pixels at intervals of 0.2 s using custom software. Since we also included calibration data for the other exercises done jointly as part the MeganePro project, a total of 498 acquisitions were processed in this manner.

The quality of eye tracking is often quantified in terms of accuracy and precision [29, 41, 7]. For each axis, the former measures the systematic error, that is, the mean offset between the actual and expected gaze locations. Precision, on the other hand, measures the dispersion around the gaze position and thus the random error of the gaze point. In Figure 2 these values are visualized with respect to the location within the video frame. This separation is intentional, as the eye tracking appears to be more accurate and precise in the center of the frame, namely (−3.5 ± 19.4) px and (−1.5± 29.6) px on the *x* and *y* axes. Moving away from the center, the gaze results systematically shift towards the borders of the frame and its random error increases. We only visualize regions where we acquired at least 40 validation samples. Pooling all data, the overall accuracy and precision is (−0.8 ± 25.8) px and (−9.9 ± 33.6) px on the horizontal and vertical axes. At a typical manipulation distance of 0.8 m, this corresponds to a real-world precision and accuracy of approximately (−0.4 ± 11.5) mm and (−4.4 ± 14.9) mm. This is deemed sufficiently accurate considering the size of the household objects used in our experimental protocol.

**Figure 2:**
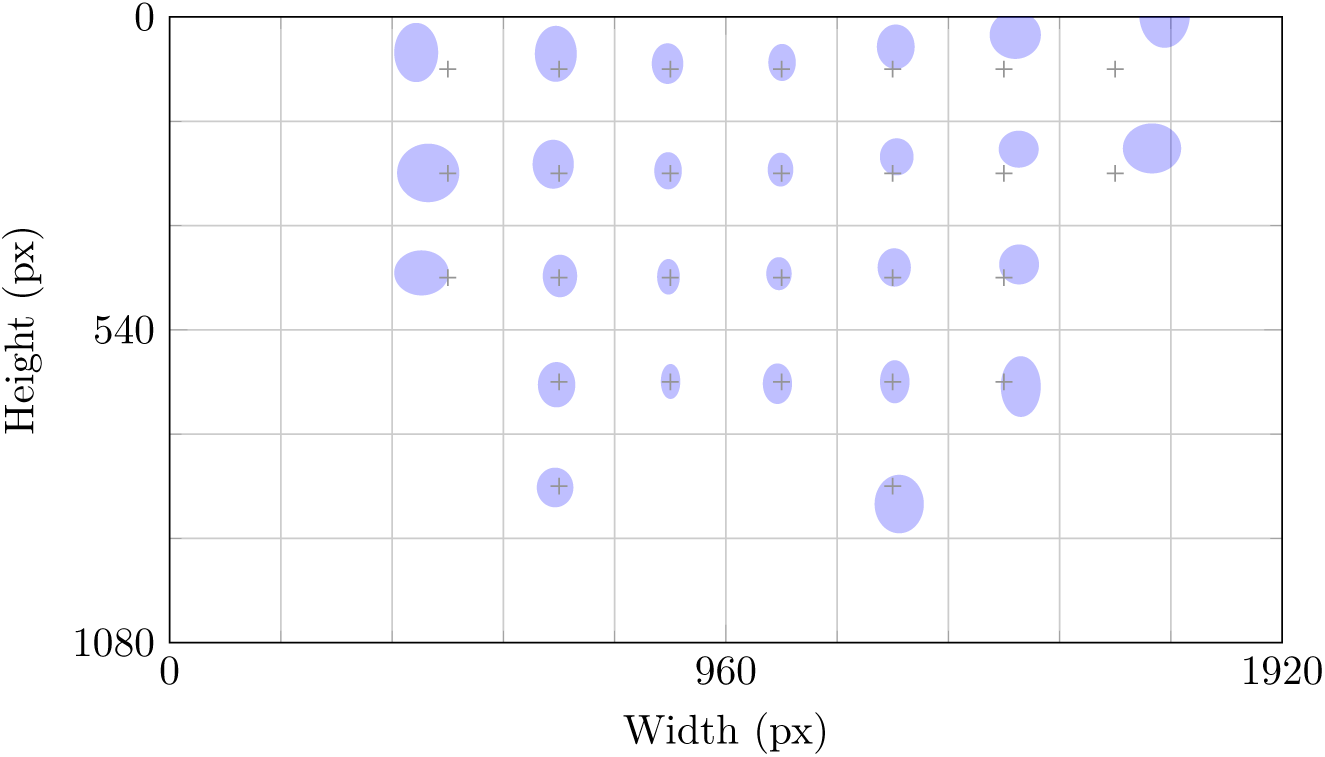
The accuracy and precision of the eye tracking with respect to the location within the video frame. For each patch, the shift of the ellipse center with respect to the cross indicates the accuracy in either axis of the gaze within that patch. The radii of the ellipse on the other hand indicates the precision.

To establish whether the calibration deteriorated over time, we compared the accuracy and precision collected at the beginning of an acquisition with those taken at the end. In total, we considered 210 uninterrupted acquisitions in which there was a calibration validation routine both at the beginning and the end. We found no statistically significant difference in accuracy (sign test, *p* = 0.95 and *p* = 1.0 in the horizontal and vertical axes) or precision (sign test, *p* = 0.24 and *p* = 0.37) indicating that drift does not pose an issue for the gaze data.

#### Statistical Parameters

To statistically describe a user’s gaze behavior during the exercises and validate it against related literature, we first identified fixations and saccades in our eye tracking data using the Identification Velocity Threshold (IVT) method [44]. To ensure that we could calculate the angular velocity of both eyes for a maximum number of samples, we linearly interpolated gaps of missing pupil data when shorter than 0.075 s [39]. We used a threshold of 70 ° /s to discriminate between fixations and saccades [34]. When the Tobii Pro glasses failed to produce a valid eye-gaze point, even after interpolating small gaps, the corresponding sample was marked as *invalid*. Excluding one subject who had strabismus, the percentage of such invalid samples ranged between (1.7 to 21.0) % and (4.3 to 30.7) % for able-bodied and amputated subjects. Sequences of events of the same type were then merged into segments identified by a time range and processed following the approach described by Komogortsev et al. [34]. First, to filter noise or other disturbances, fixations separated by a short saccadic period of less than 0.075 s and 0.5°amplitude are merged. Second, fixations shorter than 0.1 s are marked as *invalid* and excluded from the analysis.

In the resulting sequence of gaze events, the majority of invalid data are located between two periods of saccades, namely (92.2 ± 2.7) % and (92.6 ± 3.9) % for able-bodied and amputated subjects. This indicates that the Tobii Pro glasses fail predominantly to register high velocity data. Devices with sampling frequency lower than 250 Hz have indeed been categorized as “fixation pickers” [31] and often do not provide reliable results for saccades. For this reason, in the following analysis we concentrate instead on fixation events.

Figure 4 shows the distribution of the duration of fixations for both types of subjects. The characteristics of these distributions, summarized in Table 6, coincide with those described in analogous studies [30, 33, 19]. For instance, the mean values are similar to the mean duration of around 0.5 s reported by Hessels et al. [28]. Moreover, both median duration and the range between the 25^th^ and 75^th^ percentile are comparable with the results of Johansson et al. [30], who report 0.286 s as median duration and (0.197 to 0.536) s as range between the same percentiles. These similarities confirm the quality of eye tracking data and highlight that the subjects maintained a natural gaze behavior throughout the exercise.

**Table 6:**
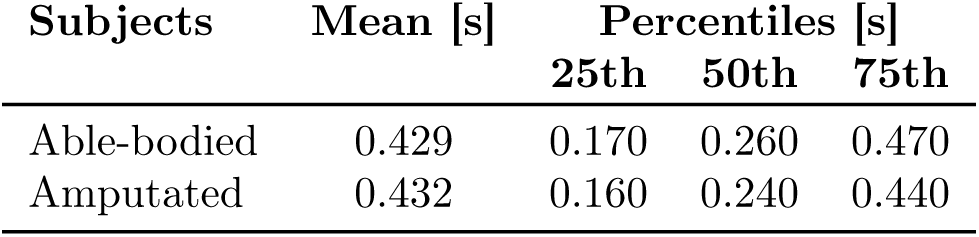
Statistical parameters of the duration of fixations for able-bodied and amputated subjects.

| Subjects | Mean [s] | Percentiles [s] |  |  |
| --- | --- | --- | --- | --- |
|  |  | 25th | 50th | 75th |
| Able-bodied | 0.429 | 0.170 | 0.260 | 0.470 |
| Amputated | 0.432 | 0.160 | 0.240 | 0.440 |

**Figure 3:**
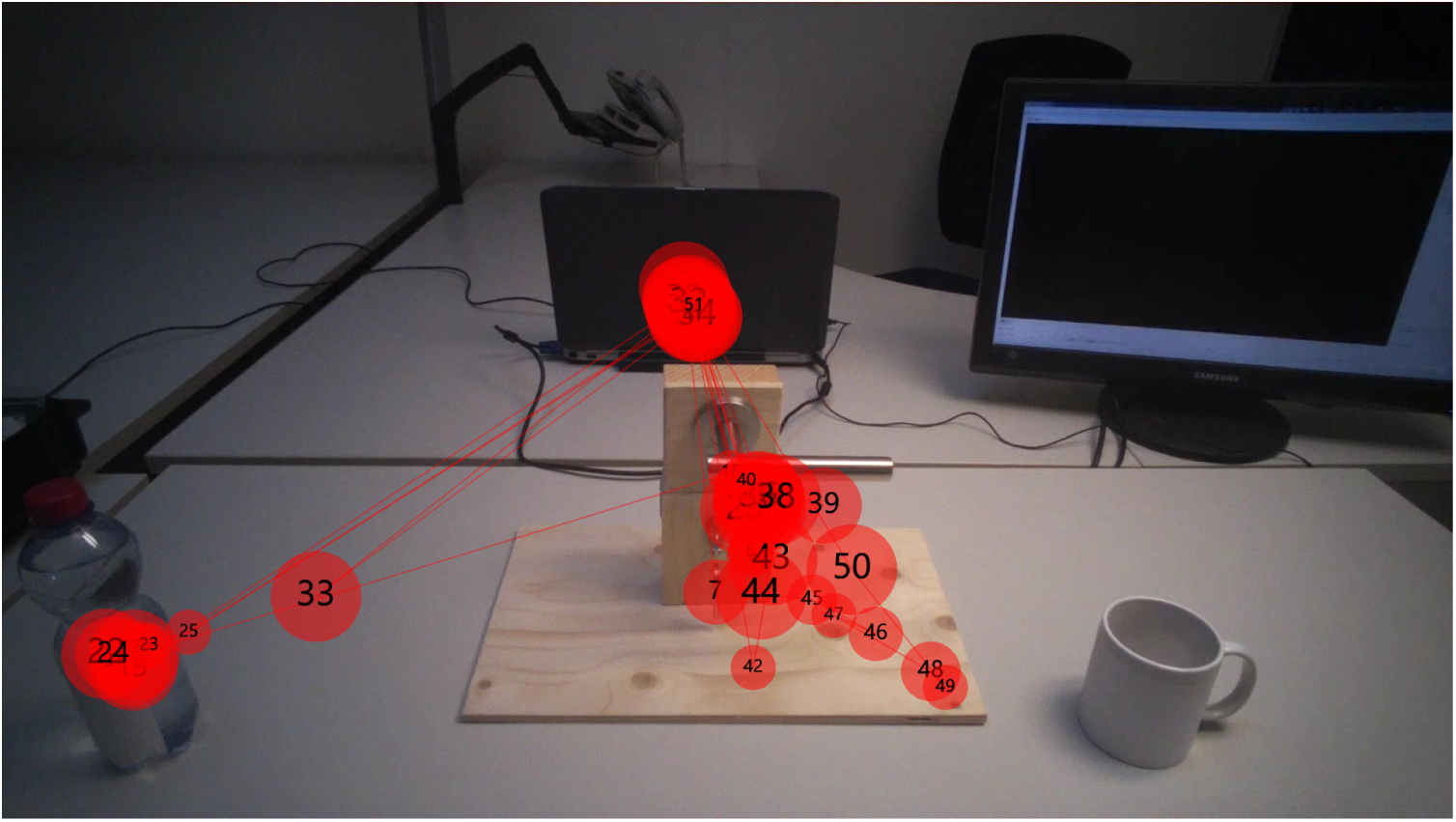
Example of gaze points overlapped onto the scene camera video. Each circle represents a fixation, where the diameter indicates the duration of each fixation and the number the order of the fixations. In this case the subject was asked to grasp the door handle and the bottle.

**Figure 4:**
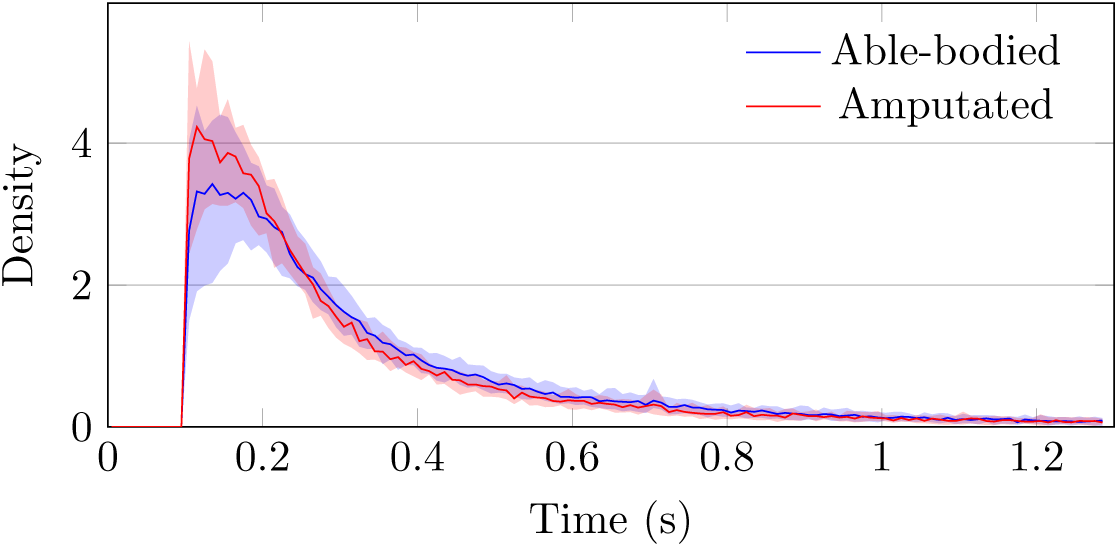
Distribution of the fixation length histogram for able-bodied (blue) and amputated (red) subjects. The shaded areas indicate the 10^th^ and 90^th^ percentiles, while the solid line represents the median.

Since one of the intended uses of these data is to investigate gaze behavior in anticipation of object manipulation, we also verified that subjects indeed looked at the object when asked to manipulate it. With the help of an automated approach to detect objects, we determined for each grasp trial whether the distance between the gaze point and the boundary of the object of interest was at least once less than 20 px. On average, this was the case in 95.9 % of the subject’s trials. Manual examination of the remaining 4.1 % indicated that the fixation exceeded the distance 20 px not because of lack of subject engagement, but due to a low accuracy calibration of the Tobii Pro glasses. Regardless, in the vast majority of the trials subjects visually located and fixated the object prior to its manipulation. For purely illustrative purposes, an example of gaze behavior of an able-bodied subject while grasping the door handle and the bottle is given in Figure 3.

### Myoelectric Signals

#### Spectral Analysis

To assure the soundness of the recorded sEMG, we first analyzed the spectral properties and compared these with known results from literature. For each subject and for each channel, we calculated the power spectral density via Welch’s method with a Hann window of length 1024 (approximately 530 ms) and 50% overlap). Figure 5a shows the distribution over these densities via its median and the range between the 10^th^ and 90^th^ percentiles. The first observation is that nearly all of the energy of the signals is contained within (0 to 400) Hz, as is typical for sEMG [6]. Furthermore, there is no sign of powerline interference at 50 Hz or its harmonics, confirming the efficacy of the filtering approach detailed in the previous section.

**Figure 5:**
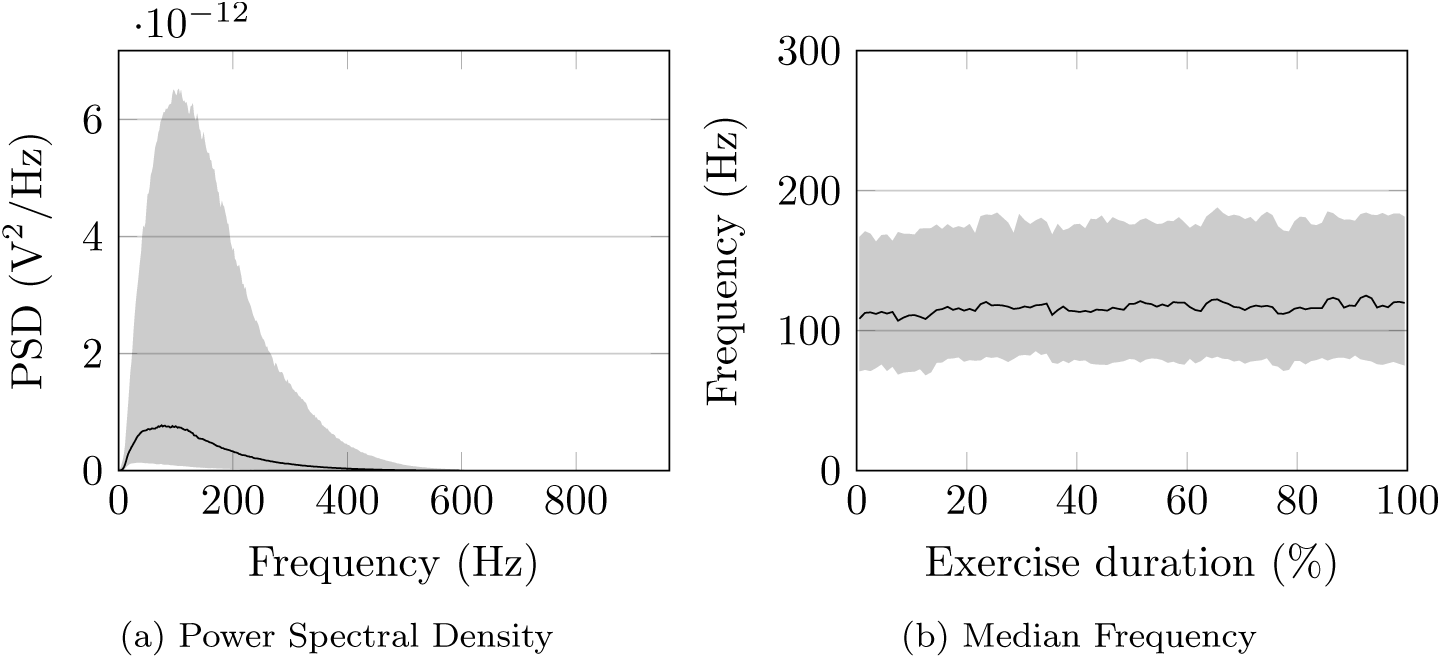
The distribution (a) of the power spectral densities and (b) the median frequency throughout the duration of the exercise. The solid line indicates the median over all subjects and electrodes, while the shaded area indicates the 10^th^ and 90^th^ percentiles.

We do however observe a rather large variability of densities among subjects and electrodes. The reason is that the spectrum and amplitude of sEMG depends on the exact position of an electrode over a muscle [37]. In our protocol, none of the electrodes was positioned precisely on a muscle belly, thus causing a wide variety in the spectrum. In some cases the signal may even be almost absent (e.g., an electrode over the radial bone).

The same variability is also noticeable in Figure 5b, which reports the distribution of the median frequency over all subjects and electrodes throughout the entire exercise. The median frequencies we find are close to the approximately (120 to 130) Hz typically reported for the flexor digitorum superficialis [11, 32], which is one on the muscles we primarily recorded from with our electrode positioning. Finally, we note that the distribution of the median frequency remains relatively stable over time, indicating that there are no persistent down- or upward shifts in the spectrum.

#### Grasp Classification

As a more high-level validation of the sEMG signals, we verify that these can indeed be used to discriminate the grasp a subject was performing, which is one of the anticipated applications of this data set. We employ the standard window-based classification approach described by Englehart and Hudgins [20] with a window length of 400 samples (approximately 208 ms) and 95% overlap between successive windows. As feature-classifier combinations we consider

- a (balanced) Linear Discriminant Analysis (LDA) classifier used with the popular four time-domain features [20];
- k-Nearest Neighbors (KNN) applied on RMS features [4]; and
- Kernel Regularized Least Squares (KRLS) with a nonlinear exponential *χ*^2^ kernel and marginal Discrete Wavelet Transform (mDWT) features [4, 26].

In contrast to prior work where we employed a single train-test split [4, 26], the classification accuracy is defined here as the average accuracy of 4-fold cross validation. In each of these folds, one of the four repetitions per grasp-object-position combination was used as held-out test data, while the remaining three repetitions formed the training data. Similarly, any hyperparameters were optimized via nested 3-fold cross validation on the train repetitions. For all methods, the training data were downsampled with a factor 10 for computational reasons, while the data used for hyperparameter optimization were downsampled with an additional factor 4.

The results in Figure 6 show a median classification accuracy between 63% and 82% for either able-bodied or amputated subjects, depending on the classification method. This is significantly higher than the approximately 50% accuracy a baseline classifier would achieve by simply predicting the most frequent *rest* class, confirming the discriminative power of the sEMG signals for the grasp type. Although a quantitative comparison with related work is of limited value due to discrepancies in experimental setup and protocol, the current results are a couple percentage points higher than those presented by Atzori et al. [4]. The most likely explanation is the lower number of grasps (i.e., only 10 rather than 40), which inevitably boosts performance. As well, for amputees, the fact that we are considering subjects with different clinical parameters from previous studies may contribute to influence the performance difference, since it has been shown that remaining forearm length, phantom limb sensation intensity, and years passed since the amputation can influence the classification outcome [5].

**Figure 6:**
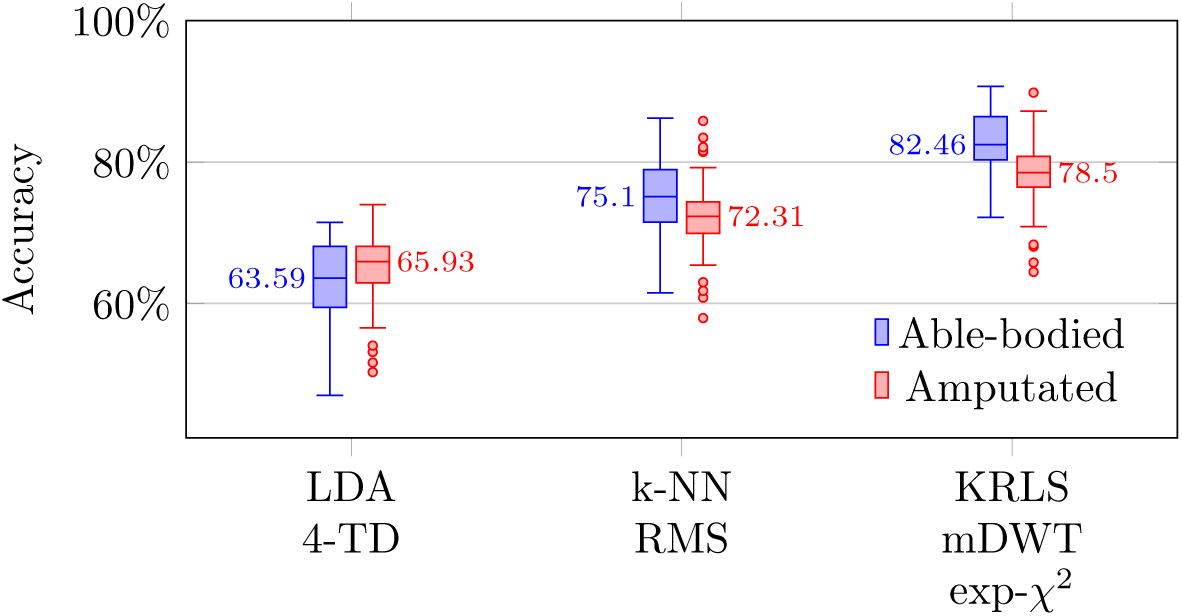
Classification accuracies for able-bodied and amputated subjects when predicting the grasp type with three different types of classifiers.

## Usage Notes

### Relabeling

The data records come with both the original stimulus as well as a variant that has been aligned to actual sEMG activation (see section Relabeling). The latter variant is preferable when the stimulus is used as ground-truth of the grasp type, such as grasp recognition, since it reduces the number of incorrect labels due to response times. In other studies one may actually be interested in these response times, such as when investigating the psychophysical response to the vocal instruction. In these cases we advise users to use the original stimulus.

### Trial Repetitions

In the experimental protocol, each new grasp started with two videos in first and third person perspective to introduce the subject to the grasp and the objects. Although subjects were encouraged to practice the grasps during this phase, some of them did not do this and focused on the video on the computer screen. For this reason the hand and gaze behavior is unreliable and users of the dataset are advised to remove movements where the (*re*)*objectrepetition* column has a value of −3 or −2.

### sEMG & Accelerometer data

A problem with sEMG and accelerometer data concerned electrode number 8 (the 8^th^ column of the *emg* variable and the 22^nd^, 23^rd^, and 24^th^ columns of the *acc* variable). Unfortunately, the data from this electrode seem unreliable. Myoelectric and accelerometer data recorded by this electrode sometimes have a substantially lower amplitude than the others, indicating a probable hardware issue. We therefore recommend to carefully consider this aspect when using the data.

### S024 & S039

Due to the difficulties reported in the previous subsection, no myoelectric data were received from electrode number 8 during these acquisitions. Therefore, the sEMG and accelerometer data for these subjects were recorded from eleven electrodes instead of twelve.

### S108

The high amputation level of S108 prevented the placement of the second array of electrodes. Therefore, only the first array consisting of eight electrodes was placed on the residual limb of this subject.

### Gaze

It is known that various factors, such as physical characteristics of the test participant, recording environment, eye tracker features, and quality of calibration affect the gaze data quality [29]. The experiment was intentionally relatively unconstrained (e.g., the head was not fixed and subjects could move their torso) to encourage natural behavior. In the following cases we have identified potential issues with gaze data that may require consideration.

### S111

A high percentage (∼ 30%) of invalid gaze data were obtained for this subject. The gaze data are included in the dataset for completeness.

### S114

For this subject, calibration could not be performed due to a strabismus condition. In such cases, the Tobii Pro glasses use a built-in default calibration. This allowed us to continue the experiment, but the quality of eye tracking is significantly worse [49]. The difficulty in tracking the subject’s eye in this particular condition is highlighted by the high proportion of invalid gaze samples (∼ 50%). Although included in the dataset, the use of the gaze data for this subject should be carefully evaluated.

### S115

A specific physical condition of this subject prevented a stable placement of the Tobii Pro glasses. The slippage compensation of the device may have helped to limit the consequences, as the proportion of invalid gaze samples (∼ 10%) indicates a proper tracking of the eye. This condition should nonetheless be taken into account when analyzing the gaze and Tobii Pro glasses’ IMU data for this subject.

### Video

MPEG transport stream (MPEG-TS) videos were recorded wirelessly via the Tobii Pro glasses Application Programming Interface (API). The variables *pts, tspipelineidx*, and *tsvideoidx* of the data records serve to synchronize the data with these videos. We however release only the MP4 videos, this for two main reasons: to limit the size of the dataset and to ensure a high quality of the scene camera videos, since the MP4 videos were immune to packet loss problems.

### Communication

Being able to select among four languages allowed the subjects to choose the one they were most comfortable with and, in most case, it corresponded to their native language. For S111 only, none of the languages available in the acquisition software were appropriate. We were however able to communicate in Italian and, in case of difficulties, communication was facilitated by a relative of the subject translating from Italian to the subject’s native language.

## Data Availability

The datasets will be released once this manuscript is accepted in a peer-reviewed scientific journal.

## Acknowledgements

This work was supported by the Swiss National Science Foundation Sinergia project #160837 “MeganePro” and by the Hasler Foundation in the project ELGAR PRO. We thank the subjects for their participation in the Myo-Electricity, Gaze And Artificial-intelligence for Neurocognitive Examination & Prosthetics (MeganePro) project. We thank Prof. G. Pajardi, Dr. E. Rosanda and the team of the hand surgery and rehabilitation department of the Ospedale San Giuseppe MultiMedica Group in Milan, Italy for their help in recruiting subjects for this study. Furthermore, we thank Mara Graziani, Francesca Giordaniello, and Francesca Palermo for their help in developing a preliminary version of the acquisition protocol and testing the acquisition software.

## Author contributions

MC created the acquisition protocol, developed the acquisition software, performed data acquisition, and wrote the paper.

AG created the acquisition protocol, developed the post-processing procedure and software, performed data analysis, and wrote the paper.

VG created the acquisition protocol, performed data analysis, and wrote the paper.

GS helped with the creation of the acquisition protocol and with the data acquisition, obtained ethical approval, and reviewed the paper.

KG recruited the participants, helped with the data acquisition, checked the videos for private data, and reviewed the paper.

A-G MH recruited the participants and reviewed the paper.

AG helped with the post-processing software, performed data analysis, and reviewed the paper.

DF recruited the participants, helped with the data acquisition and reviewed the paper.

CT recruited the participants and reviewed the paper.

FB recruited the participants and reviewed the paper.

BC ideated the project and reviewed the paper.

PB ideated the project and reviewed the paper.

MA ideated the project, created the acquisition protocol, obtained ethical approval, helped with the data acquisition, and reviewed the paper.

HM ideated the project and reviewed the paper.

## Footnotes

1 http://www.delsys.com/

2 http://www.tobiipro.com/

3 https://www.qt.io/

